# Rapid diagnosis of SARS-CoV-2 infection by detecting IgG and IgM antibodies with an immunochromatographic device: a prospective single-center study

**DOI:** 10.1101/2020.04.11.20062158

**Authors:** Felipe Pérez-García, Ramón Pérez-Tanoira, Juan Romanyk, Teresa Arroyo, Peña Gómez-Herruz, Juan Cuadros-González

## Abstract

**Objectives:** SARS-CoV-2 infection diagnosis is challenging in patients from 2-3 weeks after the onset of symptoms, due to the low positivity rate of the PCR. Serologic tests could be complementary to PCR in these situations. The aim of our study was to analyze the diagnostic performance of one serologic rapid test in COVID-19 patients.

**Methods:** We evaluated an immunochromatographic test (*AllTest COVID-19 IgG / IgM*) which detects IgG and IgM antibodies. We validated the serologic test using serum samples from 45 negative patients (group 1) and 55 patients with COVID-19 confirmed by PCR (group 2). Then, we prospectively evaluated the test in 63 patients with clinical diagnosis of pneumonia of unknown etiology that were COVID-19 negative by PCR (group 3).

**Results:** All 45 patients from group 1 were negative for the serologic test (specificity = 100%). Regarding group 2 (PCR-positive), the median time from their symptom onset until testing was 11 days. For these 55 group-2 patients, the test was positive for either IgM or IgG in 26 (overall sensitivity = 47%), and in patients tested 14 days or more after the onset of symptoms, the sensitivity was 74%. Regarding the 63 group-3 patients, median time after symptom onset was 17 days, and the test was positive in 56 (89% positivity).

**Conclusions:** Our study shows that serologic rapid tests could be used as a complement of PCR to diagnose SARS-CoV-2 infection after 14 days from the onset of symptoms and in patients with pneumonia and negative PCR for SARS-CoV-2.

## Introduction

The pandemic due to SARS-CoV-2 that started in Wuhan four months ago (1,2) has caused until April 8, 2020, a total of 1,353,361 cases and 79,235 deaths worldwide (3). Spain is the country of the European region that has been most affected by the infection, accounting for 140,510 cases and 13,798 deaths by April 8 (3). From the beginning of the pandemic, one of the main concerns was the complexity and excessive time to results of the diagnostic test, based on polymerase chain reaction (PCR) (4,5). Few clinical microbiology laboratories were prepared at this time to process such a massive volume of samples that grew exponentially. In our hospital, which is a medium-sized center (490 beds), from March 5 to April 6, a total of 7,453 respiratory samples (the vast majority nasopharyngeal exudates) were processed for SARS-CoV-2 PCR, reaching a positivity rate between 20 and 40%. Another problem was the low positivity rate of nasopharyngeal samples in patients presenting a clinical syndrome compatible with COVID-19 in the second and third week of infection (1,6-8), which is generally the period in which patients are admitted to the hospital (1). Besides, most patients presented a non-productive cough (9), and this fact, together with the high risk of generating aerosols in bronchoscopies explains that most respiratory samples came from the upper respiratory tract, where the virus concentration is lower beyond the first week after the onset of symptoms (8,10). As a consequence, the positivity rate of the PCR in these patients could be lower than expected and many of them were hospitalized with a provisional diagnosis of pneumonia of unknown etiology and possible COVID-19.

These limitations have led to development of different microplate ELISA tests (11,12). Recently published studies confirm the usefulness of combining PCR in nasopharyngeal exudates with the detection of IgM and IgG antibodies in the blood (13). The combination of molecular and serologic techniques allowed some authors to achieve a sensitivity of 97% for diagnosis of SARS-CoV-2 infection (12). However, those time-consuming tests based on ELISA are not as suitable for clinical use as rapid tests and, as a matter of fact, cannot be included in the management algorithms in emergency departments (12-15).

Since the beginning of the epidemic in Spain, information emerged about the availability of rapid serological diagnostic kits that detected IgG and IgM antibodies using immunochromatographic (ICT) tests. However, there are very few published studies about the clinical application of these kits (15). Our aim was to evaluate the diagnostic performance of one of these serologic rapid tests, first by a validation of the test in negative control patients and confirmed cases of COVID-19, and then by a prospective evaluation in patients with pneumonia of unknown etiology and a clinical diagnosis of COVID-19 with negative PCR for SARS-CoV-2.

## Methods

### Population and study period

We included three groups of patients in our study:

<u>Group 1 (healthy controls)</u>: a randomly selected group of 55 patients who had a serum sample taken for other serologic studies, from October 1 to November 30, 2019 (before the first cases of COVID-19 were reported).

<u>Group 2 (confirmed cases of SARS-CoV-2 infection)</u>: 55 patients admitted to the Emergency department between March 1 and April 6, 2020, with suspicion of COVID-19. The PCR was positive for SARS-CoV-2 for all of them.

<u>Group 3 (pneumonia of unknown etiology)</u>: 63 patients admitted for at least 5 days between February 9 and April 2, 2020, with a clinical and radiological diagnosis of pneumonia of unknown etiology, in which the PCR for SARS-CoV-2 was negative. They were prospective studied after the validation of the serologic test.

### Diagnostic methods

#### Molecular technique

Two automatic extractors were used to obtain viral RNA from clinical samples: *MagCore HF16* (RBC bioscience, Taipei, Taiwan) and *Hamilton Microlab Starlet* (Hamilton Company, Bonaduz, Switzerland). RNA amplification was made using two real-time PCR platforms: *VIASURE SARS-CoV-2 Real Time PCR Detection Kit* (Certest Biotech, Zaragoza, Spain) and *Allplex 2019-nCoV assay* (Seegene, Seoul, South Korea). All equipments were used according to the manufacturer’s instructions for both the handling and the interpretation of the results.

#### Serology

we applied the *AllTest COV-19 IgG / IgM kit* (AllTest Biotech, Hangzhou, China) for the serological diagnosis. This test is a qualitative membrane-based immunoassay (immunochromatography) for the detection of IgG and IgM antibodies against SARS-CoV-2 in whole blood, serum or plasma samples. We used 10μL of serum for the performance of the test. For the negative control group (group 1), cryopreserved archive samples were obtained, which were previously defrosted and tempered to room temperature before analysis. The performance of the test and the interpretation of the results were done according to the manufacturer’s instructions.

### Clinical data

Demographic and clinical variables of the study population were obtained from the medical records (age and sex). The time from the onset of symptoms was calculated in groups 2 and 3 from the day of onset of symptoms to the day of the extraction of the sample of serum.

### Serologic test validation

The serologic test was evaluated on clinical samples from groups 1 and 2 in order to assess the sensitivity and specificity of the test:

<u>Group 1 (negative controls):</u> they were used to evaluate the specificity of the serological test. 45 aliquots of cryopreserved sera, corresponding to 45 different controls, were recovered from the serum archive.

<u>Group 2: (patients with positive PCR for SARS-CoV-2):</u> they were used to evaluate the sensitivity of the serological test, using PCR as a gold standard. A total of 55 confirmed cases of SARS-CoV-2 infection were included, and cryopreserved aliquots of serum of those patients were used. Those aliquots were previously obtained from samples sent to the laboratory to carry out other serologies.

### Diagnostic performance of the serologic test

The assessment was performed on patients from group 3 (<u>pneumonia of unknown etiology with negative PCR for SARS-CoV-2</u>). Fresh serum samples from 63 patients were studied.

### Statistical analysis

We considered a positive result for samples in which IgG, IgM or both of them were detected. Continuous variables are expressed as median and interquartile range (IQR) and categorical variables as proportions. Comparisons between categorical variables were made using the Chi-squared or Fisher’s exact two-tailed test and the Mantel-Haenszel test for linear trends. For these comparisons, a *p* value less than or equal to 0.05 was considered significant. Statistical analysis was performed with SPSS v20.0 (IBM Corp., Armonk, NY, USA).

## Results

A total of 163 patients were studied. Median age was 62 years (IQR: 51-74); 107 (65.6%) were males. The overall serologic results from the three groups of patients are summarized in **Table 1**.

**Table 1.**
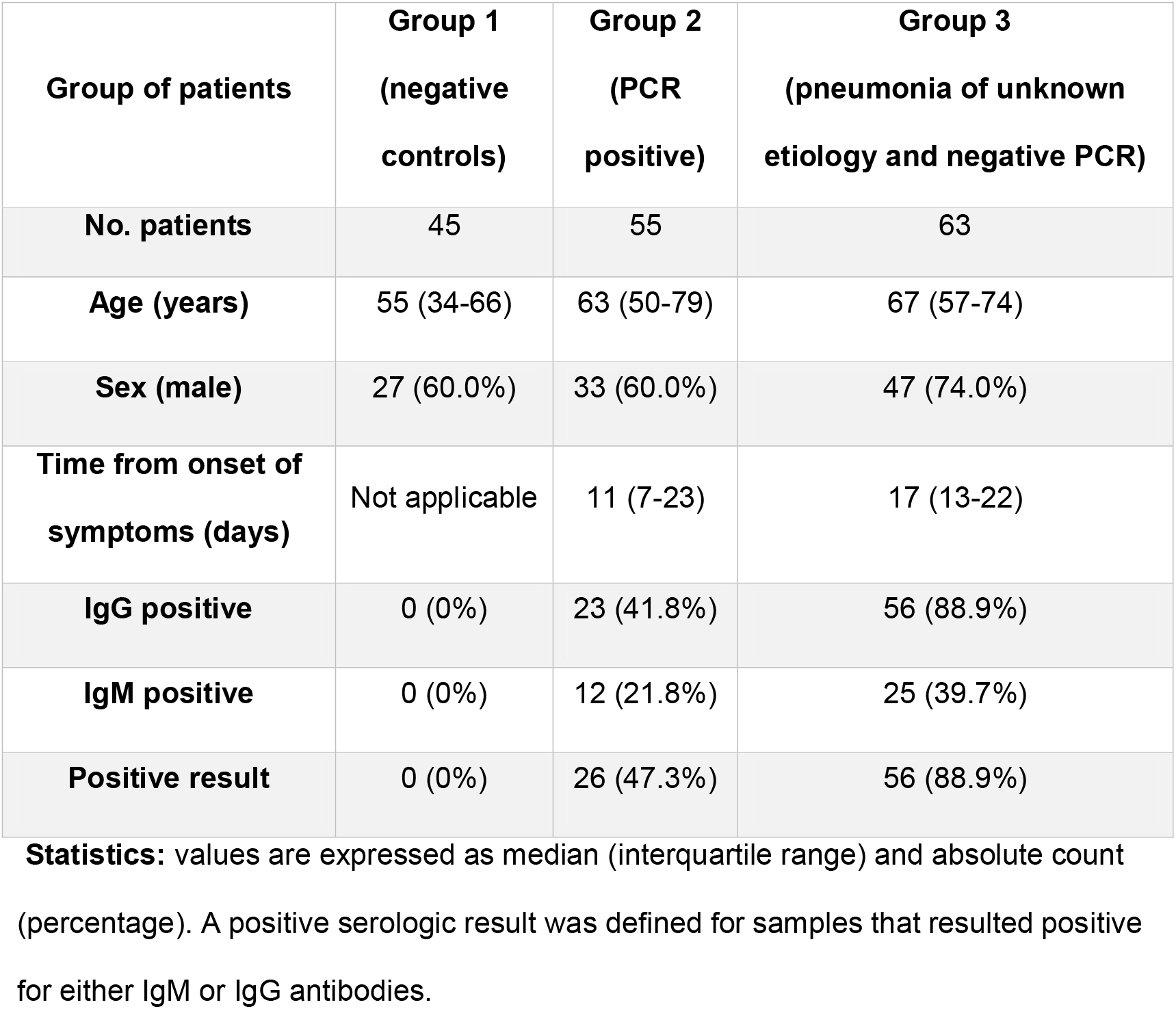
Overall serologic results from the three groups of patients.

| **Group of patients** | **Group 1**<br>**(negative**<br>**controls)** | **Group 2**<br>**(PCR**<br>**positive)** | **Group 3**<br>**(pneumonia of unknown**<br>**etiology and negative PCR)** |
| --- | --- | --- | --- |
| **No. patients** | 45 | 55 | 63 |
| **Age (years)** | 55 (34-66) | 63 (50-79) | 67 (57-74) |
| **Sex (male)** | 27 (60.0%) | 33 (60.0%) | 47 (74.0%) |
| **Time from onset of**<br>**symptoms (days)** | Not applicable | 11 (7-23) | 17 (13-22) |
| **IgG positive** | 0 (0%) | 23 (41.8%) | 56 (88.9%) |
| **IgM positive** | 0 (0%) | 12 (21.8%) | 25 (39.7%) |
| **Positive result** | 0 (0%) | 26 (47.3%) | 56 (88.9%) |

**Statistics:** values are expressed as median (interquartile range) and absolute count (percentage). A positive serologic result was defined for samples that resulted positive for either IgM or IgG antibodies.

### Serologic test validation

All patients included in group 1 (negative controls) showed negative results for serological tests. Thus, the serological test presented a specificity of 100%. The overall sensitivity of the test was 47.3% compared to PCR (**Table 1**). The sensitivity increased within the first 2 weeks both for IgM and IgG (p=0.008 and p<0.001, respectively, **Table 2**), reaching a sensitivity of 73.9% after 14 days from the onset of symptoms. **Figure 1** shows the evolution in the positivity rates of the test in group 2: for IgM antibodies, the positivity increased to a maximum level of 66.7% that was reached approximately 13 to 18 days after the onset of symptoms and then began to decrease until reaching its minimum at 31 to 36 days. IgG positivity rates increased up to 100% at 31 to 36 days after the onset of symptoms.

**Table 2.**
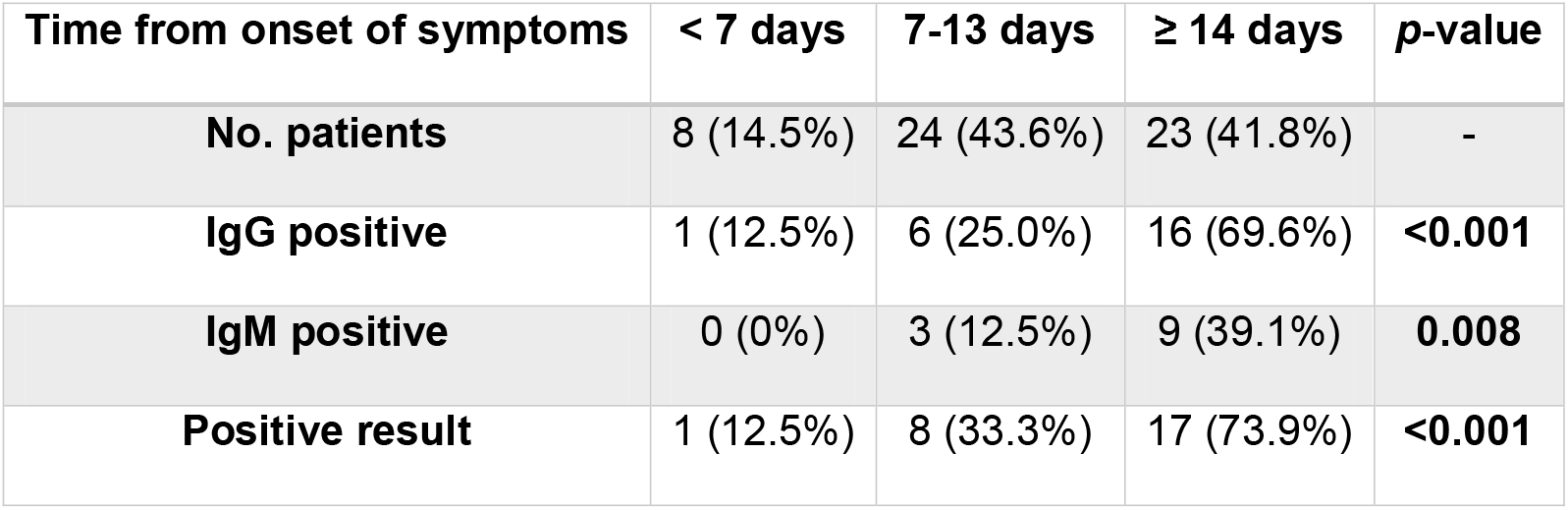
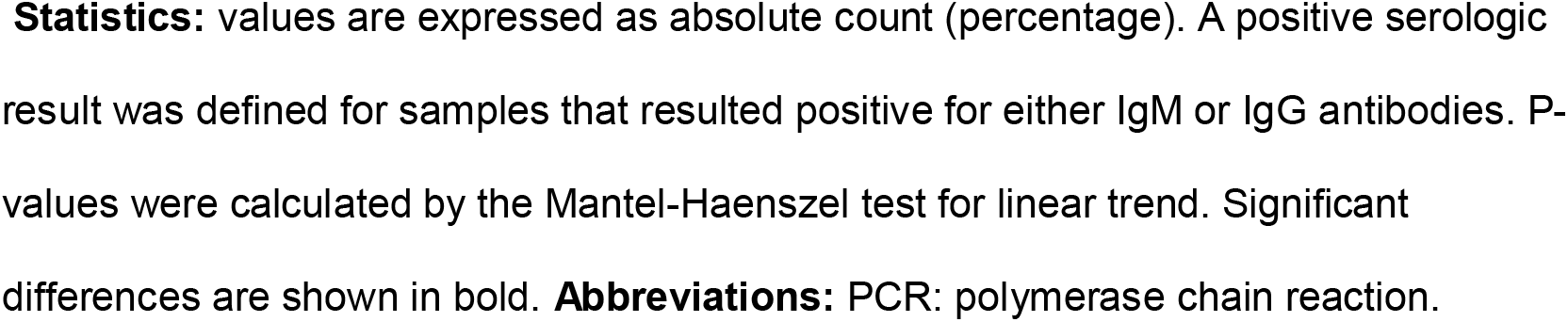
Serologic results in group 2 patients (PCR positive) according to the time from the onset of symptoms.

| **Time from onset of symptoms** | **< 7 days** | **7-13 days** | **≥ 14 days** | **p-value** |
| --- | --- | --- | --- | --- |
| **No. patients** | 8 (14.5%) | 24 (43.6%) | 23 (41.8%) | - |
| **IgG positive** | 1 (12.5%) | 6 (25.0%) | 16 (69.6%) | **<0.001** |
| **IgM positive** | 0 (0%) | 3 (12.5%) | 9 (39.1%) | **0.008** |
| **Positive result** | 1 (12.5%) | 8 (33.3%) | 17 (73.9%) | **<0.001** |

**Statistics:** values are expressed as absolute count (percentage). A positive serologic result was defined for samples that resulted positive for either IgM or IgG antibodies. P-values were calculated by the Mantel-Haenszel test for linear trend. Significant differences are shown in bold. **Abbreviations:** PCR: polymerase chain reaction.

**Figure 1.**
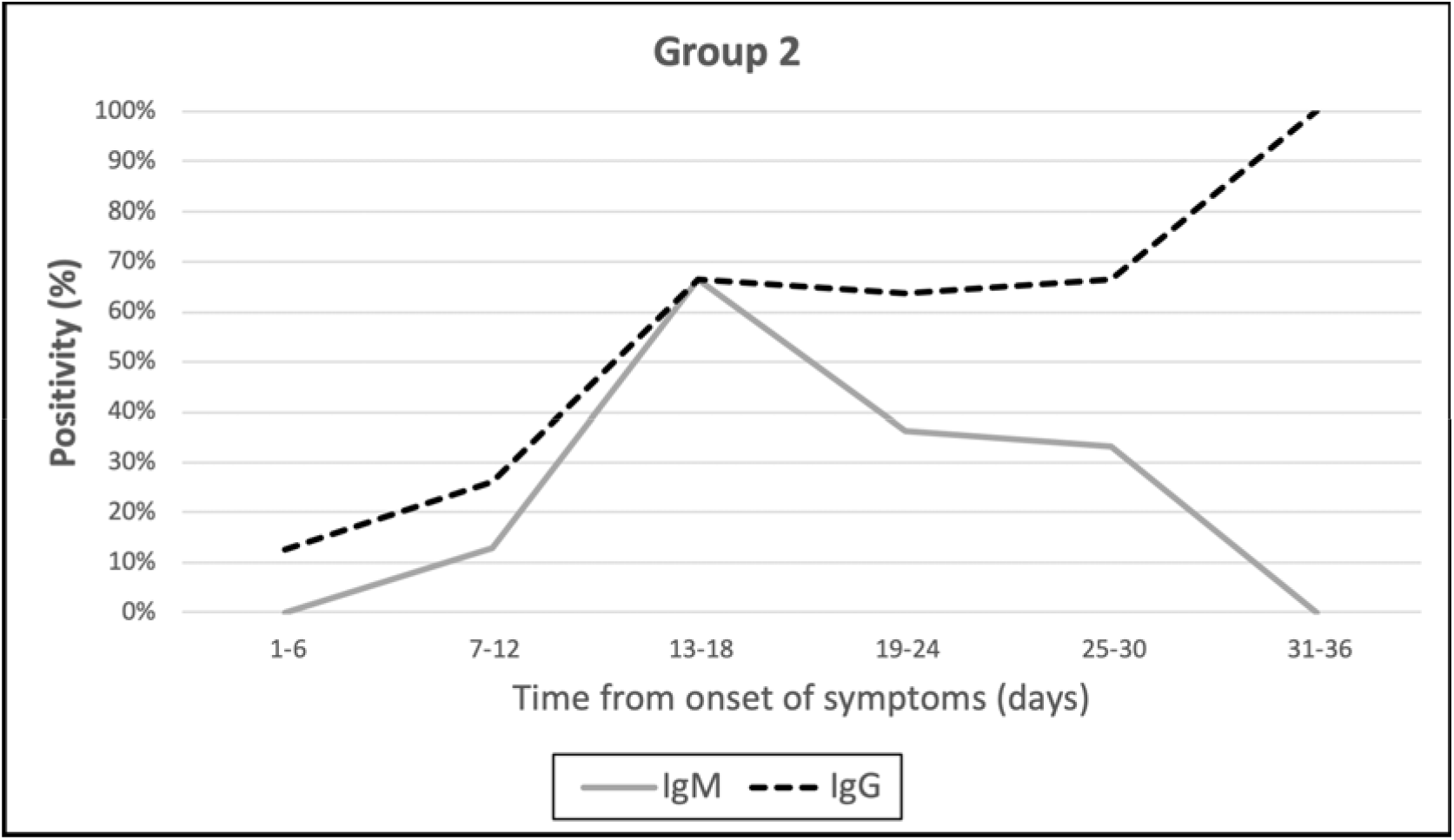
Temporal evolution of the percentage of positivity for IgM and IgG antibodies in group 2 patients (PCR positive)

### Diagnostic performance of the serologic test

We assessed the serologic test in the group 3 patients (patients with pneumonia of unknown etiology and negative PCR). Antibodies against SARS-CoV-2 were detected in 56 out of 63 patients (88.9%), being all of them positive for IgG antibodies. Twenty-five patients (39.7%) were also positive for IgM antibodies (**Table 1**). No patient had less than 7 days from onset of symptoms and the positivity rate increased from 7-13 days (83.3%) to ≥14 days (91.1%), but this difference was not statistically significant for IgM or IgG (p=1.000 and p=0.397, respectively, **Table 3**). We observed a similar pattern in the evolution of the percentages of positivity for IgM in these patients compared with those belonging to group 2 (**Figure 2**). However, group 3 patients exhibited a sustained higher positivity rate for IgG from days 13 to 30 after the onset of symptoms.

**Table 3.**
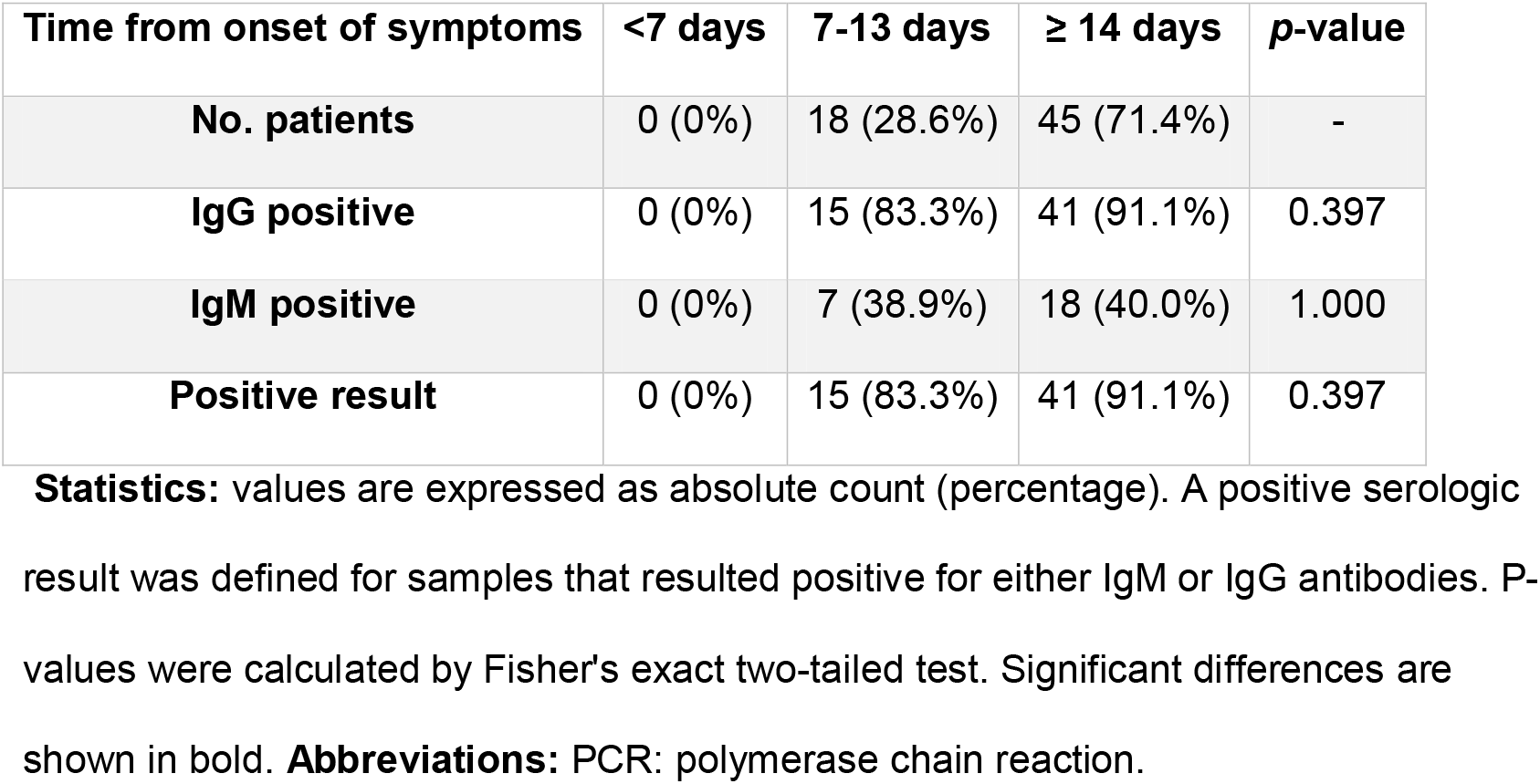
Serologic results in group 3 patients (pneumonia of unknown etiology and negative PCR) according to the time from the onset of symptoms.

| Time from onset of symptoms | <7 days | 7-13 days | ≥ 14 days | p-value |
| --- | --- | --- | --- | --- |
| **No. patients** | 0 (0%) | 18 (28.6%) | 45 (71.4%) | - |
| **IgG positive** | 0 (0%) | 15 (83.3%) | 41 (91.1%) | 0.397 |
| **IgM positive** | 0 (0%) | 7 (38.9%) | 18 (40.0%) | 1.000 |
| **Positive result** | 0 (0%) | 15 (83.3%) | 41 (91.1%) | 0.397 |

**Statistics:** values are expressed as absolute count (percentage). A positive serologic result was defined for samples that resulted positive for either IgM or IgG antibodies. P-values were calculated by Fisher's exact two-tailed test. Significant differences are shown in bold. **Abbreviations:** PCR: polymerase chain reaction.

**Figure 2.**
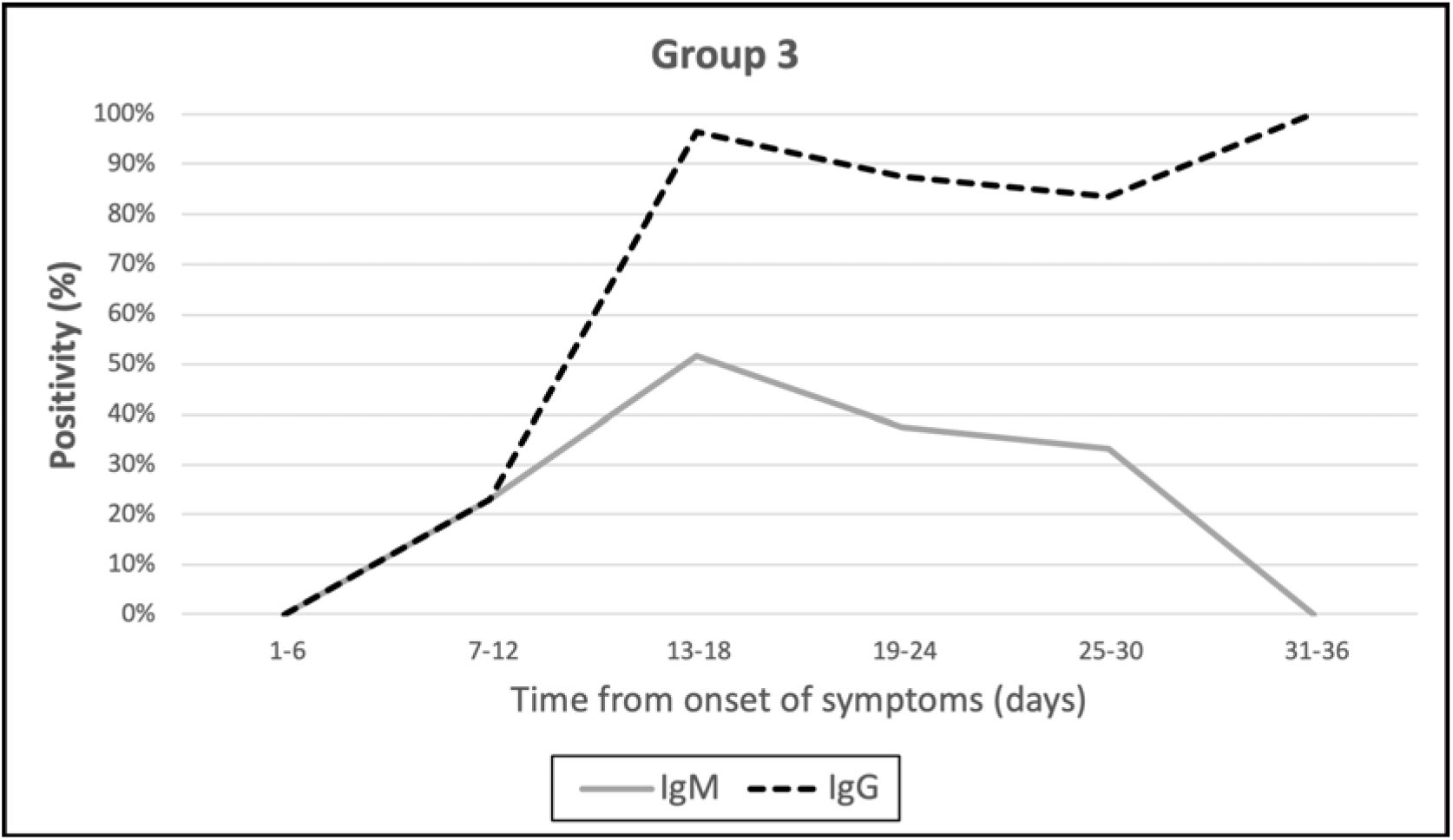
Temporal evolution of the percentage of positivity for IgM and IgG antibodies in group 3 patients (pneumonia of unknown etiology with negative PCR)

## Discussion

Our study shows that immunochromatographic tests are a reliable tool to diagnose SARS-CoV-2 infection from 14 days of onset of symptoms and are especially useful in hospitalized patients with pneumonia of unknown etiology with 14 or more days from the onset of symptoms and in whom the PCR has been negative.

The current situation of the COVID-19 pandemic requires an urgent and coordinated answer to the inherent problems of the PCR-based diagnosis: on the one hand the low capacity to carry out PCR techniques in some laboratories and also the low sensitivity of PCR test in nasopharyngeal samples, specially from the second week of infection (2,6). This study shows that the *AllTest COVID-19 IgG / IgM* rapid test for the detection of IgG and IgM is very specific (100%) and reaches a sensitivity of 73.9% from day 14 of onset of symptoms in patients with previous positive PCR in a nasopharyngeal exudate. There is increasing evidence on the usefulness of serology for diagnosis of SARS-CoV-2 infection, but most of these studies are based on microplate ELISA tests to detect IgA, IgM and IgG antibodies (12,13). These techniques have shown high sensitivity and specificity, but they also require special equipment, trained personnel and take several hours to perform. Due to this, there is an increasing interest about the usefulness of serologic rapid tests, but there is scarce information about their diagnostic performance. In a recently published study, Liu *et al*. (10) performed a multicenter evaluation of a serologic rapid test that the authors had developed. In their study, the overall sensitivity was 88.7% and the specificity was 90.6%. However, although they achieved a higher sensitivity than that obtained in our study, these authors did not present any data about the time after the onset of symptoms except from 58 out of 525 patients enrolled in the study. Moreover, for this subgroup of patients they only described that the time from the onset of symptoms was 8 to 33 days. Maybe there was a selection bias in the enrolled patients and most of the recruited cases had long evolution times, possibly leading to these results in sensitivity. To the best of our knowledge, our study is the first evaluation of this serologic rapid test which has complete data on the time from the onset of symptoms. As this is a serological test, this kind of information is key in order to interpret properly the sensitivity and specificity results. Additionally, in our experience, the use of these tests allowed diagnosis of COVID-19 infection in 91.1% of a group of 63 patients admitted with a clinical diagnosis of COVID-19 pneumonia and negative PCR in nasopharyngeal exudate. According to our data, the vast majority of patients seroconvert from day 21 and this is a key aspect in the management of health care personnel (16) and in population immunity studies related to pandemic control (17).

Our study is subject to some limitations. First, it has been conducted in a single hospital. Further multicenter studies are necessary to reinforce our findings. Second, patient selection was made according to the diagnostic needs of our hospital. Consequently, group 3 patients were all patients with negative PCR patients with clinical and radiological criteria of pneumonia and because of that, our results could not be generalized to other patients with COVID-19 and other clinical syndromes. Additionally, group 3 patients also presented a longer evolution time than group 2 patients. This probably explains that the overall positivity rates of the serological test are better than in group 3 (88.9% vs 47.3% in group 2). However, when we focus on patients with 14 or more days from onset of symptoms, the sensitivity and positivity rate increased for groups (91.1% for group 3 and 73.9% for group 2 patients). Because all of these limitations, further studies including all kinds of clinical presentations are needed in order to reinforce our conclusions.

The question about the reliability of serologic rapid tests is still under debate (18,19) and more research is needed on this topic. We think that our study may help to point out the usefulness of these rapid tests.

## Data Availability

Data are available from the authors upon reasonable request

## Acknowledgements

We want to thank Carolyn Brimley Norris, from the University of Helsinki Language Services for her help with the preparation of the manuscript.

## Compliance with Ethical Standards

### Informed consent

Since the present study is retrospective, informed consent was not required.

### Ethical approval

The study was conducted according to the ethical requirements established by the Declaration of Helsinki. The Ethics Committee of Hospital Universitario Príncipe de Asturias (Madrid) approved the study.

### Author contributions

Study concept and design: FPG, RPT and JCG

Patients’ selection and clinical data acquisition: FPG, RPT, JR, TA, PGH and JCG

Sample processing: JR, TA and PGH

Statistical analysis and interpretation of data: FPG and RPT

Writing of the manuscript: FPG, RPT, JR and JCG

Critical revision of the manuscript for relevant intellectual content: JR, JCG

Supervision and visualization: JCG

All authors read and approved the final manuscript.

